# Determinants of Digital Health Pre-Acceptance in Respiratory Care: A Quantitative Irish Study

**DOI:** 10.64898/2026.11.06.26367410

**Authors:** T. Byrne, N. Murray, M.C Donnell-Naughton, N.J. Rowan

## Abstract

**Introduction:** Digital health technology (DHT) is increasingly being integrated into healthcare delivery; however, limited research exists exploring the pre-acceptability of DHT among ageing respiratory patients. This study aimed to identify the factors influencing the pre-acceptability of digital health technology in individuals with chronic respiratory disease in Ireland.

**Methods:** A quantitative study was conducted among respiratory service users attending two respiratory centres in the Midlands and West of Ireland. A convenience-based questionnaire was developed and distributed, which was informed by prior Phase 1 qualitative findings that adopted the Technology Acceptance Model and the Health Belief Model.Data were analysed using non-parametric statistical methods, including the Mann–Whitney U test, to explore differences between gender and geographical location (urban/rural).

**Results:** Fifty-eight participants completed the study. Overall, participants demonstrated positive perceptions toward the use of DHT, particularly regarding support for self-management, hospital admission avoidance, increased symptom awareness, and enhanced engagement with healthcare professionals. Perceived usefulness emerged as a significant factor influencing acceptability. Concerns regarding privacy, data security, and digital literacy were less significant than anticipated. Statistical differences were identified between urban and rural participants in relation to perceived usefulness, technology design preferences, feedback functionality, and preference for reduction in face-to-face appointments. No significant differences were identified between male and female participants.

**Conclusions:** This study demonstrates that ageing respiratory patients in Ireland are receptive to the integration of digital health technology provided that technologies are perceived as useful, user-friendly, and supportive rather than replacements for healthcare professionals. The findings highlight the importance of involving service users during the design phase of DHT and recognising geographical differences in perspectives toward technology adoption. These findings may inform future policy, service development, and digital health implementation strategies nationally and internationally.

## Introduction

The global healthcare sector has witnessed accelerating interest in digital technologies to support and enhance clinical service delivery. The COVID-19 pandemic served as a catalyst, rapidly exposing both the potential and necessity of remote healthcare solutions. Digital health, encompassing e-health, m-health, and telehealth, offers opportunities to strengthen clinical engagement through advanced monitoring capabilities aimed at improving patient outcomes [1]. These technologies enable healthcare professionals to make more informed clinical decisions and elevate care quality, predicated on the assumption that service users possess both the capability and willingness to engage with such innovations. Consequently, digital health technologies (DHT) represent a promising paradigm for symptom monitoring, treatment adherence support, and enhanced healthcare access across diverse patient populations [2].

### Relevance to Respiratory Care

For respiratory conditions including chronic obstructive pulmonary disease (COPD) and asthma, DHT present particularly compelling opportunities to advance disease management and enable personalised care pathways [3]. The clinical evaluation schedule for this patient cohort, typically involving assessments at six-to-twelve-month intervals, may contribute to under-recognition of disease progression. While increasing consultation frequency could address this gap, practical constraints including cost, time, and geographical accessibility often limit healthcare access. Digital health technologies offer a potential solution by facilitating remote symptom self-management and ongoing monitoring.

### Digital Literacy and Adoption

Müller et al. [4] established that successful DHT utilisation is intrinsically linked to internet access and digital competence. These authors observed higher DHT adoption rates among younger, more educated, and higher-income populations, suggesting that effective engagement requires concurrent health literacy and information technology proficiency. Supporting this perspective, Neter and Brainin [5] documented that individuals with limited e-health literacy demonstrate reduced capacity to utilise available online health resources. Conversely, Smith [6] challenged assumptions regarding older adults’ technological capabilities, arguing that media portrayals have perpetuated misconceptions about this demographic’s willingness and ability to adopt technology. Perrin and Duggan [7] further contested this narrative, identifying older adults as one of the fastest-growing populations in terms of internet engagement. These conflicting perspectives underscore a critical research question: “What factors influence successful DHT adoption among ageing populations?”

### Evidence from Clinical Trials

Clinical investigations have examined DHT applications across various healthcare contexts. Steel et al. [8] evaluated remote patient monitoring for asthmatic patients aged 17-50, reporting mean compliance of 80%. Notably, 96% of participants indicated willingness to reuse the equipment and rated it as easy or very easy to use. During the study period, 48% received medical intervention, suggesting remote monitoring’s potential to reduce asthma morbidity. Ierodiakonou et al. [9] explored remote care for COPD and congestive heart failure, identifying significant reductions in emergency department presentations. While numerous studies have evaluated purpose-designed devices for chronic diseases, highlighting age, gender, and physiological measurement influences, the specific factors affecting service user engagement or disengagement remain unclear from existing research.

### The Adoption Gap

Global DHT adoption patterns reveal significant variation, with wearable technologies lagging considerably behind established technologies such as smartphones and tablets [10]. Cheung et al. [11] identified a concerning knowledge gap regarding service users’ adoption intentions in healthcare contexts. Much existing research has prioritised technical development over understanding diffusion processes, contrasting sharply with commercial technology marketing research, which focuses predominantly on consumer adoption and user experience, thereby accelerating diffusion. Lycett et al. [12] advocated for integrating psychological theories— including social cognitive theory, the health belief model, and self-efficacy theory—into digital intervention development to enhance effectiveness and adoption rates.

### Acceptance and Sustained Engagement

Nadal et al. [13] highlighted that user acceptance varies across pre-acceptance, initial acceptance, and sustained use phases, each presenting distinct design challenges that influence compliance and long-term engagement. The literature reveals considerable definitional variability regarding technology acceptance. Perski and Short [14] conceptualised acceptability as an individual’s attitude or willingness to engage with novel therapies or digital devices, encompassing pre-engagement willingness, actual usage, and post-engagement satisfaction. Recent recommendations by Nadal et al. [15] identified a gap in understanding pre-acceptance of technologies and emphasised the need to capture user perspectives post-usage. While post-usage evaluation may assess effectiveness, it fails to illuminate factors necessary for sustained engagement. As healthcare continues its digital evolution, understanding service user perceptions becomes imperative.

### Study Aim

This study therefore aims to identify and describe the factors influencing DHT pre-acceptability among respiratory patients as a specific end-user population.

## Materials and Methods

Quantitative research is often referred to as a “confirmatory” or “deductive” approach as its main goal is to test theories and hypotheses by examining the relationships among variables [16, 17]. These variables in turn can be measured so that numbered data can be analysed using statistical procedures [17]. The questionnaire was developed not only using qualitative data from phase 1 which was previously published [18], but also by the amalgamation of the Technology Acceptance Model and the Health Belief Model. The Technology Acceptance Model was developed in the 1980s in an attempt to help researchers explore the rationale as to why individuals were not using technology that was made available to them [19]. The use of validated frameworks in conjunction with the data collected from phase 1 enhanced the rigor of the questionnaire, and ultimately resulted in a study that was well-founded. External persons from the study were acquired for face and validity testing who reviewed the developed questionnaire independently.

### Participants

The participants for the second phase of the study were recruited in two different Institutions by means of purposeful sampling. A cohort of respiratory patients attending Portiuncula University Hospital and Galway University Hospital were recruited from 02/02/2022 to 13/04/2022. They were invited to complete a questionnaire. Written information was provided to each participant; consent was obtained and assurance that their decision not to participate would not have any impact on the care provided at that clinic.

### Inclusion and Exclusion Criteria

Participants in this study were women and men resident in Ireland. Inclusion criteria included service users with known respiratory disease attending respiratory services in the midlands/west of Ireland, English-speaking, and ≥60 years of age. Exclusion criteria included non-English speaking individuals and those on active palliative pathways.

### Ethics

Ethical approval is important for all types of research. This is to ensure the protection of participants, the researcher and the organisation they represent. It aims to minimise the risk of harm to all involved. Participants were provided written information in the form of a handout about the study, its pros and cons along with the benefits of participating in the study. Ethical approval was first sought from the Technological University of the Shannon Research Ethics Committee, and upon gaining approval then ethical permission was sought from clinical sites. Applications for ethical approval contained a copy of detailed information sheets and consent forms which was also attached for consideration. The completed submissions were made on 12th December 2021 and approval was granted on January 15th, 2022. The researcher carried out data collection from March 2022 to May 2022 (Ethical approval number C.A.2734).

### Sample Size

The average number of patients attending both clinics over a 10-week period was estimated to be 300. The assumption was of an average non-attendance rate of 15% [21] and an overall expected completion rate of 33% [22], with the estimated sample size for this study being 84. The duration of this study was dictated by the secondment period agreed for the researcher and the permission of access to clinics by management in the HSE.

### Data Collection

Participants could choose to complete the questionnaire at the time of the clinic or return via a supplied stamped addressed envelope provided.

### Data Analysis

Data was collected in the form of a questionnaire utilising a Likert Scale, which is ordinal data and cannot be considered normally distributed as the values are bound on the left and right. To confirm that data were indeed not distributed normally, the researcher conducted a Shapiro Wilk Test, which did further confirm with a significance of .000 across both variables that the data was not normally distributed; therefore, for the purpose of this study non-parametric tests were considered for statistical analysis. The non-parametric test selected was the Mann-Whitney U Test.

## Results

### Study Population

This study was conducted over a ten-week period collecting data from two separate institutions in the Midland and West of Ireland, with fifty-eight (58) individuals participating. Of the fifty-eight participants, 44.8% (n=26) were male and 53.4% (n=32) were female. The mean average of those who disclosed their age was 69.5. Geographically, 32.8% (n=19) self-reported to be from an urban area and 55.2% (n=32) from a rural location. In respect to technology skills, 52% (n=30) claimed to have self-taught skills, with no prior formal education. Over 300 questionnaires were distributed with only 58 respondents, despite the provision of stamped addressed envelopes.

### Key Themes

The key themes yielded from this study were:

• Support of self-management of respiratory disease;

• Engagement with healthcare professionals;

• Design and ability of the technology to provide data feedback;

• Security;

• Digital literacy;

• Perceived usefulness and awareness of digital health technology.

### Support for Self-Management of Respiratory Disease

When participants were asked whether DHT would assist them in managing their respiratory disease, 26.5% (n=13) strongly agreed and 32.6% (n=16) agreed, while 32.6% (n=16) were unsure and 4% (n=2) disagreed. Regarding hospital admission avoidance, 18.5% (n=10) strongly agreed and 37% (n=20) agreed that DHT would help reduce admissions, with 38.2% (n=21) uncertain and 5.5% (n=3) in disagreement. Concerning feelings of security between healthcare appointments, 8.4% (n=4) strongly agreed and 46.9% (n=23) agreed that DHT would increase their sense of security, while 34.6% (n=17) were unsure and 10.2% (n=5) disagreed. Notably, 17.3% (n=9) strongly agreed and 38.4% (n=20) agreed that DHT would empower them to take control of their chronic disease, with 40.8% (n=21) uncertain and 17.3% (n=9) disagreeing.

Mann-Whitney U tests comparing male versus female and urban versus rural perspectives revealed no statistically significant differences across any self-management or admission avoidance variables (all p > 0.05).

### Engagement with Healthcare Professionals

When asked whether they derived more value from face-to-face appointments, 33.3% (n=18) strongly agreed and 37% (n=20) agreed, with 14.8% (n=8) uncertain and an equal proportion disagreeing. Regarding whether DHT would reduce required clinic appointments, 12% (n=6) strongly agreed and 48% (n=24) agreed, while 34% (n=17) were unsure and 6% (n=3) disagreed. Interestingly, 9.5% (n=4) strongly agreed and 40.4% (n=17) agreed that they would prefer a reduction in appointments, with 26.1% (n=11) uncertain and 23.8% (n=10) disagreeing.Finally, when asked if DHT would increase overall contact with healthcare providers, 10.8% (n=5) strongly agreed, 39.1% (n=18) agreed, 39.1% (n=18) were unsure, and 10.8% (n=5) disagreed. Mann-Whitney U tests demonstrated no significant differences between genders regarding any engagement variable (all p > 0.05). Similarly, no significant differences emerged between urban and rural participants regarding value placed on face-to-face clinics or perceived increased contact with healthcare professionals (p = 0.60 and p = 0.81, respectively). However, a statistically significant difference was observed between urban and rural cohorts regarding preference for reduction in face-to-face clinics (p = 0.01). Rural participants demonstrated less preference for reduced clinic attendance compared to their urban counterparts.

### Technology Design and Feedback Capabilities

Regarding technology size as a factor influencing adoption, 8.5% (n=4) strongly agreed and 36.1% (n=17) agreed that size would affect their decision, while 36.1% (n=17) were unsure and 19.1% (n=9) disagreed. Visual acuity was considered important by 4.3% (n=2) who strongly agreed and 47.8% (n=22) who agreed, with 17.39% (n=8) uncertain and 30.4% (n=14) disagreeing. Notably, 37.2% (n=19) strongly agreed and 50.9% (n=26) agreed that they would prefer the device to have the capability to provide health data feedback, with 11.7% (n=6) uncertain and no participants disagreeing.Mann-Whitney U tests revealed no significant differences between males and females regarding technology size, design features, or feedback capabilities (p > 0.05). Conversely, statistically significant differences emerged between urban and rural participants regarding design features (p = 0.01) and the ability to provide health information feedback (p = 0.01). Rural participants demonstrated less favourable attitudes toward DHT design features and expressed less interest in feedback capabilities compared to urban participants. No significant difference was observed regarding the impact of technology size (p = 0.08).

### Security and Privacy

When asked whether DHT would invade their privacy, 7.3% (n=3) strongly agreed, 29.2% (n=12) were unsure, and 63.4% (n=26) disagreed. Regarding concerns about who would have access to their health data, 14.6% (n=6) strongly agreed, 21.9% (n=9) agreed, 21.9% (n=9) were unsure, and 41.4% (n=17) disagreed. Concerning technology security, 17.02% (n=8) strongly agreed, 10.6% (n=5) agreed, 31.9% (n=15) were unsure, and 40.02% (n=19) disagreed. Mann-Whitney U tests confirmed no statistically significant differences across gender or geographical variables (p > 0.05).

### Digital Literacy

When asked to consider the statement “I do not have the IT skills to use technology,” 17.6% (n=9) strongly disagreed and 41.1% (n=21) disagreed, while 15.6% (n=8) strongly agreed, 9.8% (n=5) agreed, and 15.6% (n=8) were unsure. Mann-Whitney U tests revealed no significant differences between genders (p = 0.43) or geographical locations (p = 0.46).

### Perceived Usefulness

When presented with the statement “Digital health technology is something that I feel would be useful to me,” 18.8% (n=10) strongly agreed, 49% (n=26) agreed, 30% (n=16) were unsure, and 1.8% (n=1) disagreed. Mann-Whitney U tests demonstrated no significant difference between male and female perspectives (p = 0.52). However, a statistically significant difference emerged between urban and rural cohorts (p = 0.04), with rural participants expressing fewer positive perceptions of DHT usefulness.

### Awareness of Digital Health Technology

When asked to consider the statement “I have never heard of digital health technology prior to participating in this study,” 23.9% (n=11) strongly agreed, 30.4% (n=14) agreed, 30.4% (n=14) disagreed, and 15.2% (n=7) were unsure. Mann-Whitney U tests confirmed no significant differences between genders (p = 0.47) or geographical cohorts (p = 0.23).

## Discussion

### Perceived Usefulness

Perceived usefulness is one of the most important factors discussed in the literature when discussing acceptability and DHT. People’s understanding of why they are using DHT and what benefits it has for them significantly influences adoption. This is evident in studies such as Shinoda et al. [10] whose research shows that wearable technology is accepted when it facilitates goal setting, provides reminders with useful feedback, and has the ability to share data with HCPs. Cerdán de las Heras et al. [29], Hamine et al. [30] and Shinoda et al. [10] identify that the acceptability of technology is profoundly influenced by the service users’ belief in why they are using the technology, what benefits it will have for them and how easy the technology is to use. In this study, most participants strongly agreed or agreed that DHT would be useful to them. Statistically, males and females’ perspectives do not differ (p=0.528); however, there is an evident statistical difference between urban and rural cohorts (p=0.048). There is a lack of clarity as to why there is a statistical difference between those from rural and urban areas. One possible reason is simply a lack of awareness. As previously discussed, the literature shows no evidence of exploration of the awareness of DHT. This study found that 54.3% of participants had never heard of DHT prior to the study. While there was no statistical difference between males and females or urban and rural areas, it is most definitely a concern. There is an expectation or even worse, an assumption by those developing technology and HCPs that service users will engage with DHT. Service users need to understand clearly why they need to use DHT and what benefits it will have for them. Future research into the perspectives of urban and rural geographical location is required to gain a deeper understanding. Those involved in the development of DHT, policy makers and clinicians need to be aware that there is a statistical difference, and remain cautious if assuming what works for urban areas will work for rural areas.

### Digital Literacy

Digital literacy is defined as a person’s ability to find, evaluate, and communicate via a technology device. Digital health literacy is the ability to find, understand, and appraise health information from electronic sources and apply the knowledge gained to address or solve health problems [5]. The literature clearly shows that empowering service users to engage with DHT is heavily dependent on their digital literacy skills [4, 28]. In the same studies, digital literacy was reported to be a significant adoption barrier; often individuals did not possess any digital literacy skills, resulting in an inability to interpret and act upon the readings of the technology device.Contrary to the argument made by Neter and Brainin [5], Smith [6] is confident that communities such as the elderly fall victim to misconceptions concerning their use of technology. Perrin and Duggan [7] discredit this and commend the elderly population as being one of the fastest growing populations in engaging in internet use. Participants in this study were asked to consider the statement “I do not have the IT skills to use technology.” A majority (58.7%) strongly disagreed or disagreed with this statement. There were no statistical differences between males and females or those from rural and urban areas. Digital literacy skills are an important contributing factor to the pre-acceptability of DHT, and this study shows that a majority of participants feel they have the skills required to engage.

### Data Privacy and Security

The concepts of privacy and accessibility of personal health data were addressed with service users. When asked if they felt DHT would invade their privacy, 63.4% (n=26) disagreed. When asked if they had concerns as to who would have access to their health data, 41.4% (n=17) disagreed. Finally, when asked if they would have concerns about the security of the technology, 40.02% (n=19) disagreed. There was no statistical difference between the variables measured and the null hypothesis was retained. The perspectives of participants in this study appear contrary to other published studies [25, 32]. While it is important to safeguard the privacy and health data of service users, this was not a major concern for the ageing respiratory patients and, therefore, it can be argued that data privacy is not a significant contributing factor to the pre-acceptability of DHT in this cohort.

### Support of Self-Management of Respiratory Disease

Self-management of chronic respiratory diseases is widely discussed in the literature and is strongly advocated for within appropriate guidelines [33]. There is evidence to suggest that DHT increases patients’ awareness of their condition, reduces hospital admissions, and improves communication with healthcare professionals [12]. Slevin et al. [28] and Korpershoek et al. [25] report that participants viewed DHT as extremely useful in helping them to independently manage their chronic disease.This study found that 59.1% of participants agreed or strongly agreed that DHT would assist them in managing their respiratory disease, and 55.5% agreed it would help reduce hospital admissions. Additionally, 55.7% agreed that DHT would empower them to take control of their chronic disease. There was no noted statistical difference between perspectives of males and females or those from urban and rural locations. These findings have the potential to have a massive impact on the healthcare system. The growth of the ageing population in the Republic of Ireland has accelerated compared to other EU countries. In 2019, the estimated population of individuals greater than 65 was 696,300 (14% of total population), estimated to reach 1.6 million by 2051 [36]. The number of patients waiting for a Respiratory Consultant appointment is estimated to be 19,200 [37]. The use of DHT has the potential to support self-management, reduce the need for hospital admission, and create awareness of symptoms prompting early medical advice.

### Engagement with Healthcare Professionals

Participants were asked if they felt that they get more value from face-to-face appointments with HCPs, if they felt DHT would reduce the number of face-to-face clinics required, if they would prefer a reduction in clinic appointments, and if they felt DHT would give them more contact with their HCPs. The majority (70.3%) strongly agreed or agreed that they get more value from face-to-face clinics. When asked if DHT would reduce the amount of clinic appointments required, 60% strongly agreed or agreed.Interestingly, 49.9% strongly agreed or agreed that they would prefer a reduction in appointments, with 23.8% disagreeing. This cohort appears to value face-to-face appointments, in line with findings from the Netherlands [25]. There is a lack of clarity around the rationale; one reason may be an overall lack of awareness of DHT. Finally, when asked if DHT would increase overall contact with their HCP, 49.9% strongly agreed or agreed.There are no differences in perspectives between males and females or those from different geographical regions regarding value on face-to-face clinics and contact with HCPs. However, there is a noted statistical difference (p=0.017) among those from urban and rural locations on their preference for a reduction in face-to-face clinics.

### Design and Ability to Provide Data Feedback

Nadal et al. [13] explain that our perceptions of technology evolve over time, and people may take up a new device only to discontinue its use a week later. User acceptance is frequently overlooked at the design phase, even though it is essential that the design accounts for the users’ experience from pre-acceptability to sustained acceptability [13].

This study aimed to address this gap by exploring preferences regarding technology design and capabilities. Results showed that 44.6% agreed that technology size would influence their decision, 52.1% agreed visual acuity was important, and an overwhelming 88.1% agreed they would prefer the device to have the ability to feedback their health data. It is clear that design and functionality are important contributing factors to the pre-acceptability of DHT. Statistically, there are differences between the variables evaluated. While males and females share similar perspectives, there appears to be a statistical difference between urban and rural areas regarding design features (p=0.018) and the ability to provide health information feedback (p=0.016). It is imperative this difference is noted and taken into consideration during the design phase of DHT.

### Implications for Policy

There is an urgency both at National and International level to develop policy documents in retrospect of the introduction of DHT. Findings from this study will advance Digital Health Strategies in Ireland and Europe. While digital health policies are in their infancy, successive National policies have used limited retrospective enquiry from patient perspective. From an overarching system or national policy perspective, this use of bottom-up patient perspective will provide critical insight into understanding and eventual compliance with important new policies.Indicative outputs from this study indicate the following factors may be considered to inform key performance indicators: reliability, ease of use, numbers of patients supported expressing satisfaction, reduction in negative health outcomes, numbers up-taking new technology, cost savings to healthcare, adoption by healthcare service providers trans-regionally and internationally, policies informed, and adoption into education and training. To date there are no KPIs identified for the measurement of DHT, ultimately there are no measurements of risk.

### Implications for Future Research

This study highlights multiple areas requiring future research. Firstly, there is an apparent difference in perspectives of people from urban and rural geographical locations. There is a lack of clarity on the reasoning behind these differences, which needs further investigation. It is imperative that technology design is founded on research findings indicative of what service users want. There will be potential exclusion of cohorts if these differences are not addressed.

One concerning outcome arising from the literature is that only one study addressed sustainability. Shinoda et al. [10] reported 90% compliance at initiation dropping to 40% by week twelve. Research exploring sustainability of DHT is essential. An understanding into what motivates Irish people to continue to engage with technology is required.

Risk measurement and risk assessment are to the fore of any healthcare intervention; however, there is a heightened awareness when referencing DHT. None of the studies in the scoping review referred to risk. There were no quantifiable measures of risk identified nor explanation of protective strategies embedded in the study protocols. There is an urgency both at National and International level to develop such documents.

Finally, to date, there is no evidence of standardisation of digital health technology. In healthcare, the introduction of a new medication is processed via a rigorous protocol supported by research data. There is a significant lack of policy and protocol to standardise the use of DHT in Ireland at present. This is most definitely an area in need of urgent investigation.

## Conclusion

This study is novel and the first to explore the pre-acceptability of digital health technology in the ageing respiratory patient. This study shows that there is overall a lack of awareness of digital health technology and as a result poor trust. It is evident that respiratory patients are indeed very open minded and willing to embrace the use of DHT. A majority felt that they have the skills to engage and seemed open minded. There were some differences noted in the perspectives of those from urban and rural geographical locations, and the rationale for this is somewhat unclear.The results indicate strong viewpoints of service users’ preferences for what they would like DHT to look like and what functionality they would like it to have. There is a positive role for DHT in the management of chronic disease, and participants do believe its use has merit pending the design. Service users would also need encouragement that DHT is not intended to replace the HCP but to complement a service. The skills required to use the technology and the overall security of the data were less of a concern than anticipated for this cohort.It is hoped that the findings of this study will contribute to policy and KPI development for digital health technology on a national and international level. If consideration is given to the factors that contribute to the pre-acceptability of DHT, then future technology and implementation into practice will be based on a foundation of fact, not assumption..

## Abbreviations

DHT: Digital Health Technology
HCP: Healthcare Professional
COPD: Chronic Obstructive Pulmonary Disease
GDPR: General Data Protection Regulation
HSE: Health Service Executive
KPI: Key Performance Indicator

## Declarations

The authors declare that the research was conducted in the absence of any commercial or financial relationships that could be construed as a potential conflict of interest.

## Conflicts of Interest

The authors declare that the research was conducted in the absence of any commercial or financial relationships that could be construed as a potential conflict of interest.

## Ethical Approval

Ethical applications were first sent to the Technological University of the Shannon Research Ethics Committee, and thereafter the clinical sites. The completed submissions were made on 12th December 2021 and were approved via email on January 15th, 2022. The researcher carried out data collection from March 2022 to May 2022. (Ethical approval number C.A.2734)

## Consent to Participate

Not applicable.

## Consent for Publication

Not applicable.

## Availability of Data and Materials

The raw data supporting the conclusions of this article will be made available by the authors without undue reservation.

## Funding

Not applicable.

## Data Availability

Data will be fully available

## Acknowledgments

TB acknowledges funding support from the Technical University of the Shannon for this Doctorate study.

## Author Contributions

TB, NR, NM, and MM-N contributed to the ideation, methods development, and review of the first draft, proof editing and final review. TB undertook the study as a component of a doctoral degree. NR, NM, and MM-N supervised the PhD for data generation and paper synthesis. All authors contributed to the article and approved the submitted version.

## Notes

### Competing Interest Statement

The authors have declared no competing interest.

### Author Declarations

Ethics approval was granted

